# Challenges and lessons learned from the rapid operationalization of a prospective cohort to study the natural history and neurodevelopmental outcomes of postnatal Zika virus infection among infants and children in rural Guatemala

**DOI:** 10.1101/2022.05.12.22274987

**Authors:** Alejandra Paniagua-Avila, Daniel Olson, Amy Connery, D. Mirella Calvimontes, Guillermo A. Bolanos, Molly M. Lamb, Desiree Bauer, Aida Ralda, Neudy Rojop, Eduardo Barrios, Andrea Chacon, Melissa Gomez, Paola Arroyave, Sara Hernandez, Maria Alejandra Martinez, Saskia Bunge-Montes, Alison Colbert, Kareen Arias, Garret Brazeale, Andrea Holliday, Kay M. Tomashek, Hana M. El Sahly, Wendy Keitel, Flor M. Munoz, Edwin J. Asturias

## Abstract

During the course of the 2015-2017 outbreak of Zika virus (ZIKV) infection in the Americas, the emerging virus was recognized as a congenital infection that could damage the developing brain. As the Latin American ZIKV outbreak advanced, the scientific and public health community questioned if this newly recognized neurotropic flavivirus could affect the developing brain of infants and young children infected after birth. We report here the design, methods and rapid operationalization of a prospective natural history cohort study aimed at evaluating the potential neurological and neurodevelopmental effects of postnatal Zika virus infection in infants and young children, which had become epidemic in Central America. This study enrolled a cohort of 500 mothers and their infants, along with nearly 400 children 1.5-3.5 years of age who were born during the initial phase of the ZIKV epidemic in a rural area of Guatemala. Our solutions and lessons learned while tackling real-life challenges may serve as a guide to other researchers carrying out studies of emerging infectious diseases of public health priority in resource-constrained settings.

## Introduction

In 2015, an outbreak of Zika virus (ZIKV) was reported in Brazil and other Latin American countries.^1^ Within months, the World Health Organization (WHO) declared the epidemic a Public Health Emergency of International Concern, due to its association with microcephaly and other neurological disorders.^1^ ZIKV is now recognized as a congenital infection that can damage the developing brain, resulting in numerous sequalae known as Congenital Zika Syndrome (CZS).^2^ In addition to microcephaly, CZS is associated with eye abnormalities, hearing loss, seizures, and other health conditions.^3^ Even in the absence of physical abnormalities, CZS may cause abnormal neurodevelopment, similar to other congenital infections such as rubella and CMV.^3,4^ As the ZIKV outbreak advanced, the scientific and public health community asked if this newly recognized neurotropic flavivirus could also affect the developing brain if contracted by infants and young children after birth.

The National Institute of Allergy and Infectious Diseases (NIAID), through the Vaccine and Treatment and Evaluation Unit (VTEU) network supported our proposal to assess the neurological and neurodevelopmental impact of postnatal ZIKV infection in infants and young children. The Center for Human Development (CHD) of the Fundación para la Salud Integral de los Guatemaltecos (FUNSALUD), which had an ongoing maternal-child health program, “Creciendo Sanos,” had ample experience with active surveillance for dengue virus (DENV) infections and was well positioned to carry out this study.^5^ The earliest known case of ZIKV infection was reported in this region of Guatemala in April of 2015.^6,7^ Consequently, the University of Colorado (CU), Baylor College of Medicine (BCM) and CHD partnered to design and conduct this prospective natural history cohort study to evaluate the neurological and neurodevelopmental effects of postnatal Zika virus infection.

The first section of this manuscript describes the study methods and study procedures. The second section describes the challenges and lessons learned from the rapid operationalization of a large prospective cohort study in response to the ZIKV epidemic in a rural, resource-limited setting in Guatemala. Our approach to overcoming real-life challenges encountered in a resource-constrained setting may serve as a guide to other researchers carrying out future studies of ZIKV and other emerging infectious diseases of public health priority. Results from the study described in this paper will be published in a future manuscript.

## Methods

### Study objectives

The primary objective of this prospective natural history cohort study was to characterize the acute and post-acute clinical, neurological and neurodevelopmental outcomes of postnatally-acquired, laboratory-confirmed ZIKV infection in newborn infants and their mothers (infant-mother pairs), as well as older children ages 1.5 – 3.5 years old who may have been exposed to ZIKV during the 2015-2017 outbreak. The study also aimed to describe the clinical characteristics, viral load and kinetics of humoral immune responses during acute laboratory-confirmed ZIKV infection in infants and young children, including the potential for antibody-dependent enhancement (ADE) during acute ZIKV infection in those previously exposed to or with maternally-derived antibodies against DENV.

### Study Population

Children and mothers were enrolled from 22 communities, living within a 200 km^2^ area in the southwest Trifinio (SWT) region of Guatemala, a rural lowland area that borders Chiapas, Mexico and is hyperendemic for vector-borne infections **(Figure 1)**^8^. A community needs assessment conducted in 2012 identified high levels of food insecurity, poverty, poor access to healthcare, and high maternal-child morbidity and mortality in this area.^5^ In 2013, as a response to the needs assessment, CHD, an academic-private global health partnership (CU and AgroAmerica), began implementing clinical services and community maternal-child programs to register and follow pregnant women and their children under 3 years of age to improve child growth and development.^9,10^

**Fig 1.**
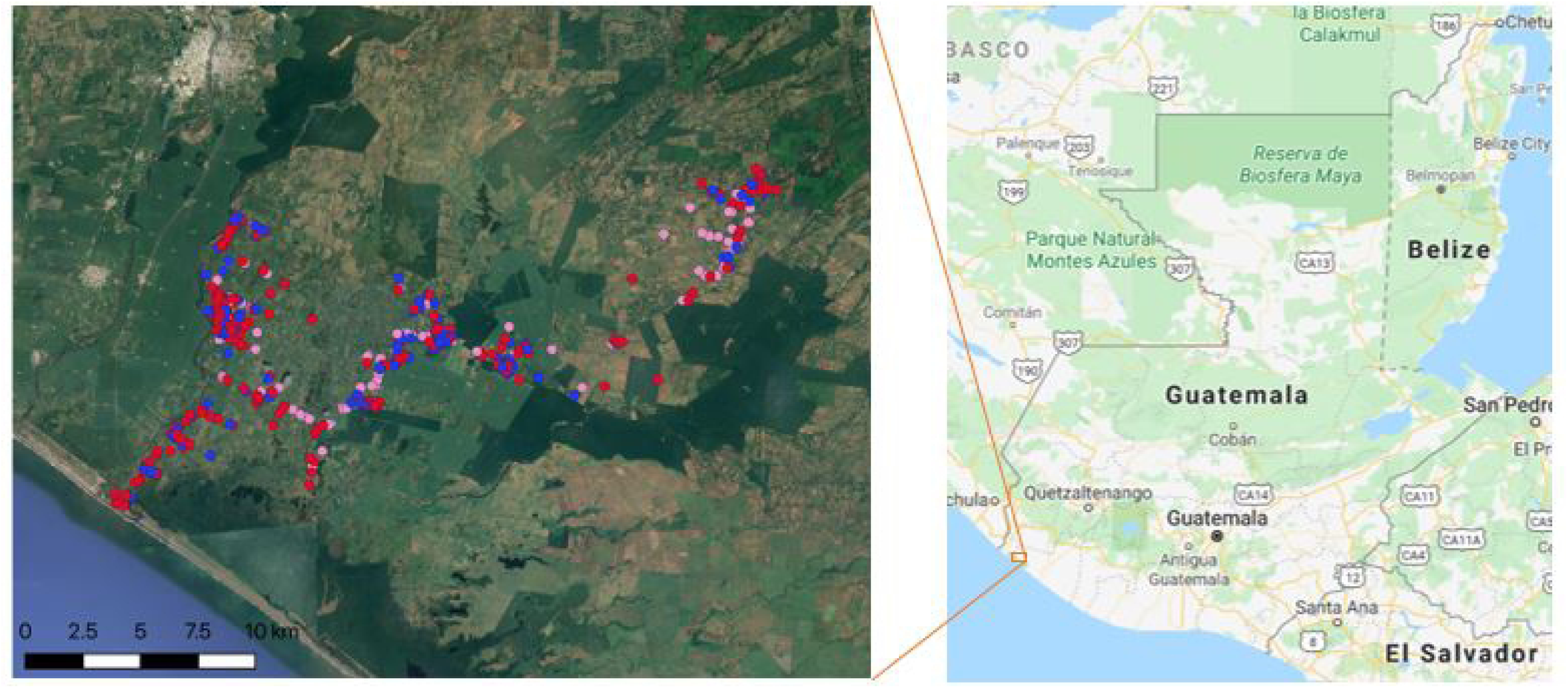
Map of study coverage area in Southwest Trifinio. This image shows the mapping points for households by study cohort.

### Eligibility criteria

Infant-mother pairs (cohorts B and M, respectively) were eligible if the child was 0-2.9 months of age, and the mother was ≥16 years of age. Informed consent was signed by the mother if ≥18 years old. If the mother was 16-17 years of age a grandparent signed the informed consent and the mother provided assent (as per local ethics committee requirements). Older children 1.5-3.5 years of age at enrollment were eligible if they either participated in a prior 2015-2016 dengue acute febrile illness (AFI) surveillance study (cohort A) or were a sibling of an infant enrolled in cohort B (cohort C), and consent for participation in this study was signed by one of the parents.

### Study Procedures

#### Participant enrollment

Infant-mother pairs were screened and enrolled from the CHD maternal and child health program and from referrals of pregnant women who were close to delivery or had delivered an infant in the previous two months by community health workers. Children 1.5-3.5 years of age who previously participated in the dengue AFI surveillance study were contacted for enrollment in this study.

#### Baseline and follow-up visits

At baseline, demographic information, including each child’s date of birth, sex, birth weight (if available), gestational age (reported by mother), the location of their home, and household characteristics were recorded. Medical history and information about initiation of breastfeeding and history of vaccinations were recorded. The child’s length, weight, and head circumference were measured using the WHO recommended procedures. ^11^ Baseline physical, neurological, neurodevelopmental, ophthalmologic, and hearing evaluations were conducted in all infants and children. For mothers, prenatal and delivery history along with past medical history were obtained, as well as measurements of mother’s height and weight. Biological samples (blood, urine, and saliva) were collected from all children, infants and their mothers. Breastmilk was also collected from mothers if available.

Protocol-required household visits at various intervals were conducted by trained field staff at the subjects’ homes or at the study clinic in order to collect health data and to perform anthropometric measurements, clinical and neurological exams, neurodevelopmental tests, and ophthalmologic and hearing evaluations. Blood, urine and breast milk samples were also collected during household visits (**Table 1)**.

**Table 1.** Study schedule of procedures for each study cohort.

| Cohort and study procedures | Study Month |  |  |  |  |
| --- | --- | --- | --- | --- | --- |
|  | 0 | 3 | 6 | 9 | 12 |
| **B – Infants - Enrollment age: 0-2.9 m** |  |  |  |  |  |
| Consent and enrollment | x |  |  |  |  |
| Neurological and Physical Exam | x | x | x | x | x |
| Hearing Test and Ophthalmological Evaluation | x |  | x |  | x |
| MSEL | x |  | x |  | x |
| ASQ3 | x | x | x | x | x |
| PedsQL | x | x | x | x | x |
| Blood | x | x | x |  | x |
| Urine | x | x | x |  | x |
| Saliva | X | X | X | X | X |
| Active FLI surveillance |  |  |  |  |  |
| **A, C – Children – Enrollment age: 1.5-3.5 y** |  |  |  |  |  |
| Consent and enrollment | X |  |  |  |  |
| Neurological and Physical Exam | X |  | X |  | X |
| Hearing Test and Ophthalmological Evaluation | X |  |  |  | X |
| MSEL | X |  | X |  | X |
| ASQ3 | X |  | X |  | X |
| PedsQL | X |  | X |  | X |
| Blood | X |  | X |  | X |
| Urine | X |  | X |  | X |
| Saliva | X |  | X |  | X |
| Active FLI surveillance |  |  |  |  |  |
| **M – Mothers** |  |  |  |  |  |
| Consent and enrollment | X |  |  |  |  |
| Physical Exam | X |  |  |  |  |
| Blood | X |  |  |  | X |
| Urine | X |  |  |  | X |
| Saliva | X |  |  |  | X |
| Breast Milk | X |  |  |  | X |
| Active FLI surveillance |  |  |  |  |  |
| MSEL – Mullen Scales of Early Learning; PedsQL – Pediatric Quality of Life Inventory; ASQ3 – Ages and Stages Questionnaire 3<sup>rd</sup> Edition; FLI – Flavivirus Like Illness |  |  |  |  |  |

#### ZIKV and DENV case ascertainment

Both ZIKV and DENV infections can be asymptomatic or subclinical in 70-80% of the infected population, or present as an acute illness with or without fever ^12,13^. Therefore, we conducted active illness surveillance using a case definition of flavivirus-like illness (FLI) which included two or more of the following signs or symptoms: fever >38.0° C for ≥ 2 days, rash, conjunctivitis (non-purulent/hyperemic), arthralgia, myalgia, or peri-articular edema for ≥ 1 day. Surveillance was conducted by weekly home visits with collection of saliva samples every other week, regardless of the presence of symptoms (**Figure 2)**. Caregivers responded to a standardized questionnaire designed to document the presence of FLI and other Integrated Management of Childhood Illness (IMCI) danger signs ^14^. If a child or mother met defined FLI criteria, blood, urine and saliva samples were collected within 72 hours of illness report and a second time at the 2^nd^, 3^rd^ or 4^th^ week after the first visit to evaluate seroconversion. The timing of the second visit was selected at random for each study subject.

**Figure 2.**
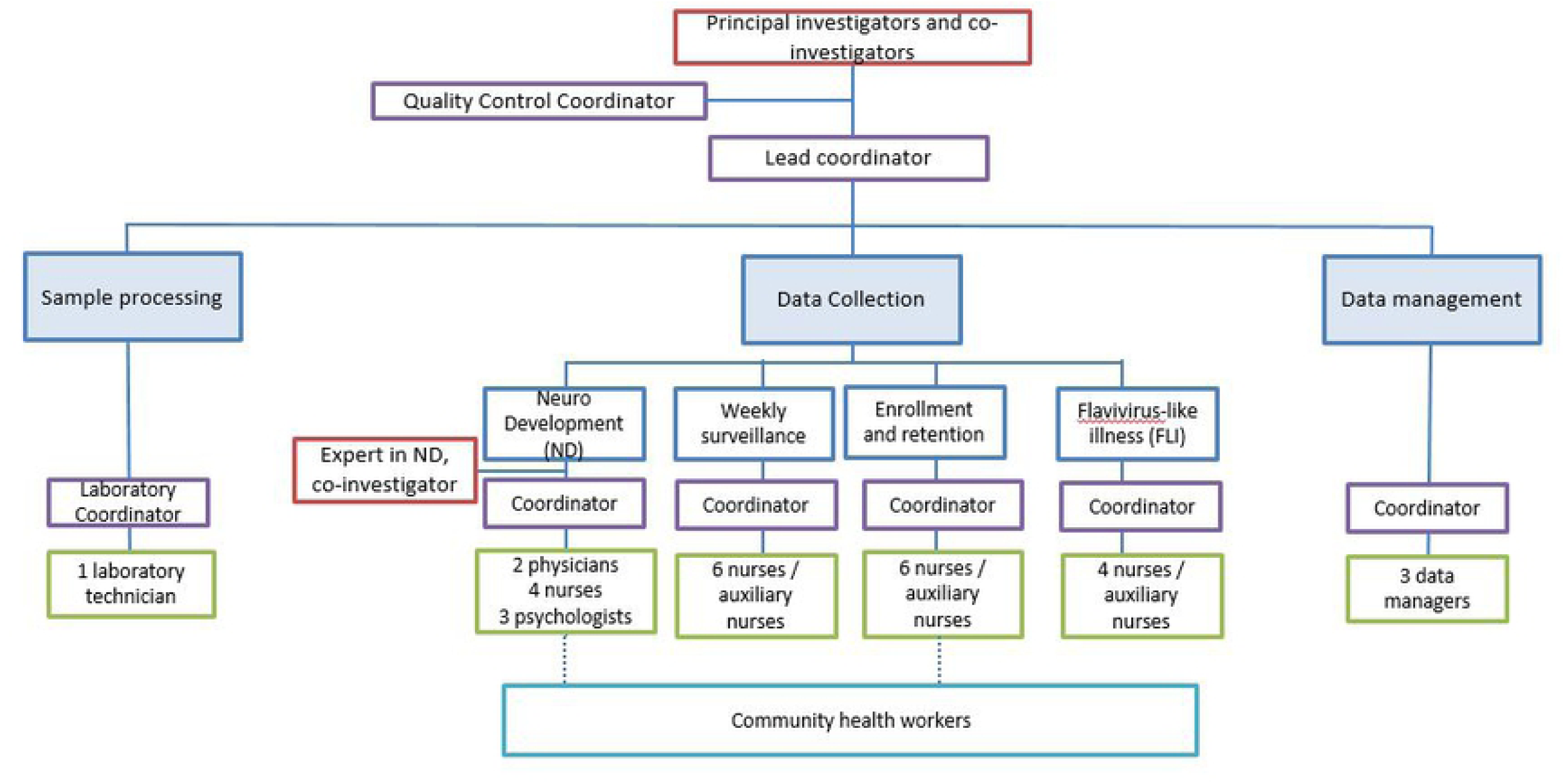
Organization of competency-based teams.

### Bioethical approval

The study was approved by the Institutional Review Boards of BCM, CU (COMIRB), and the Ministry of Public Health and Social Welfare (MSPAS) of Guatemala in May of 2017.

### Operationalization of the study protocol

A description of the challenges encountered when planning and operationalizing the study and the implemented solutions to address them are summarized in **Table 2**. Our lessons learned and recommendations outlined in **Table 3**, could guide other researchers and clinicians conducting prospective, complex studies in other low-resource settings in response to emerging infections.

**Table 2.**
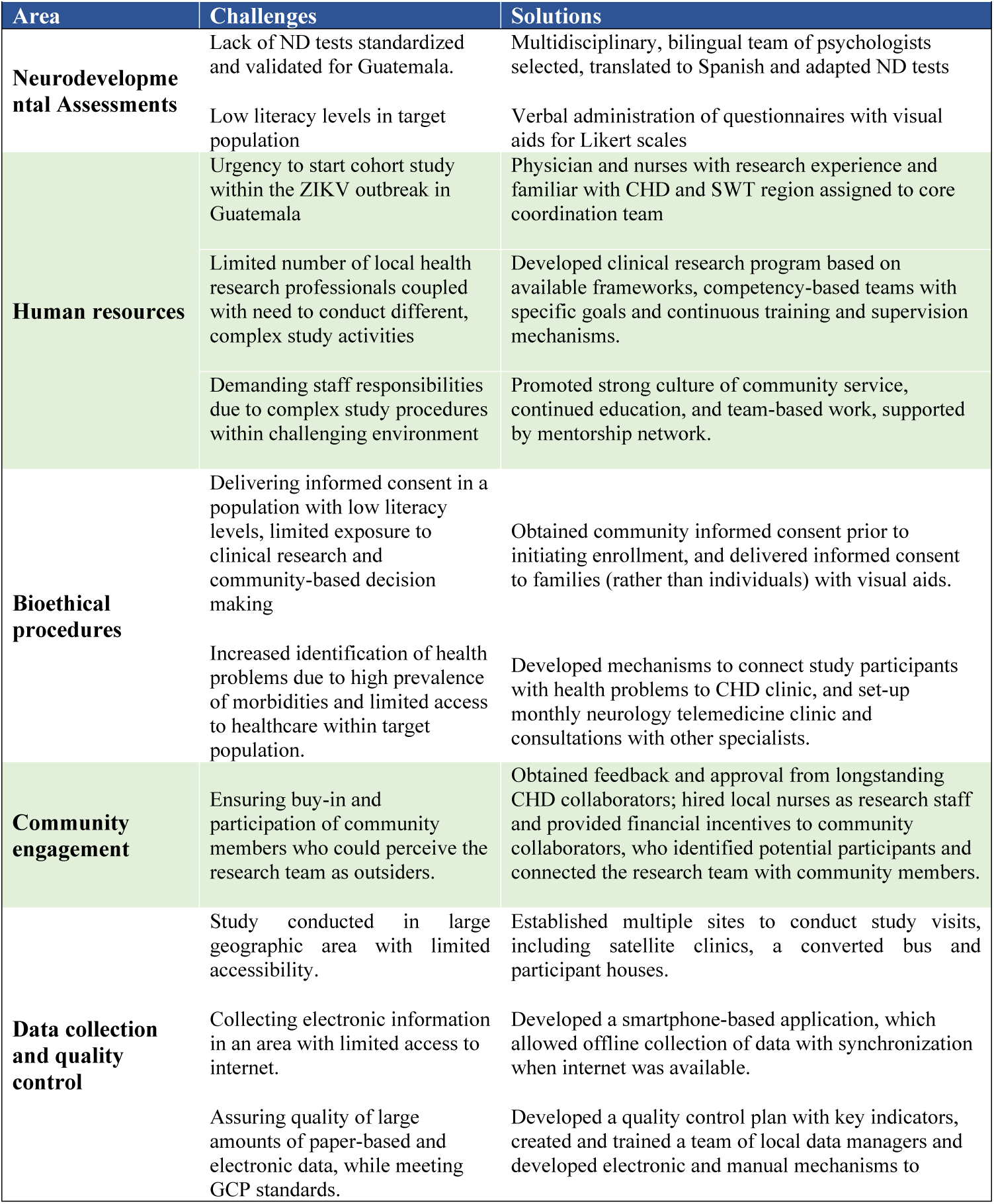
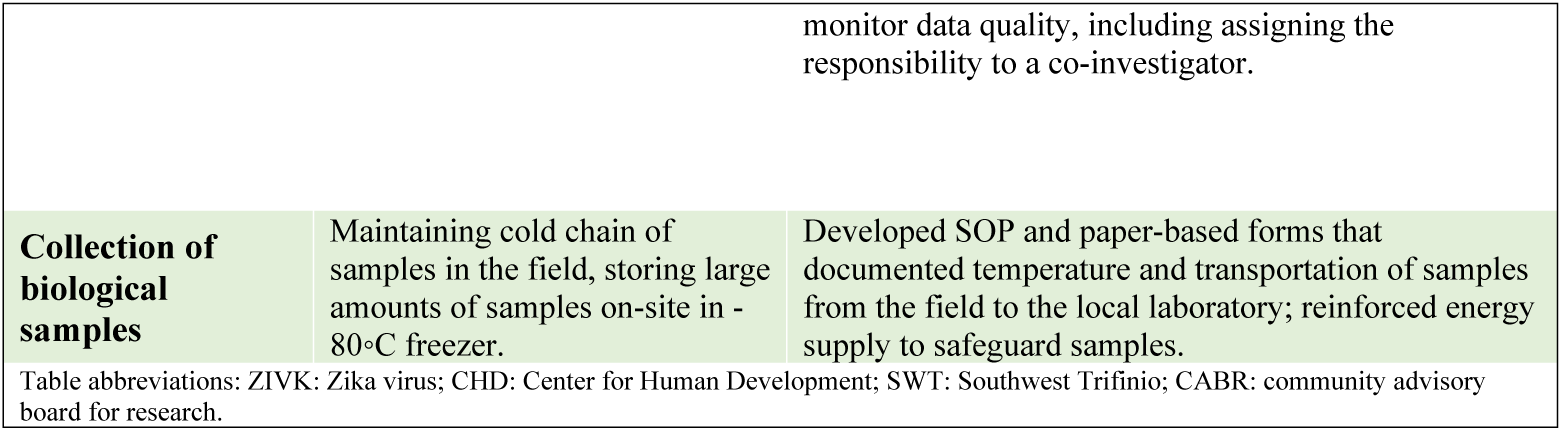
Challenges of operationalizing the study protocol and implemented solutions.

**Table 3.**
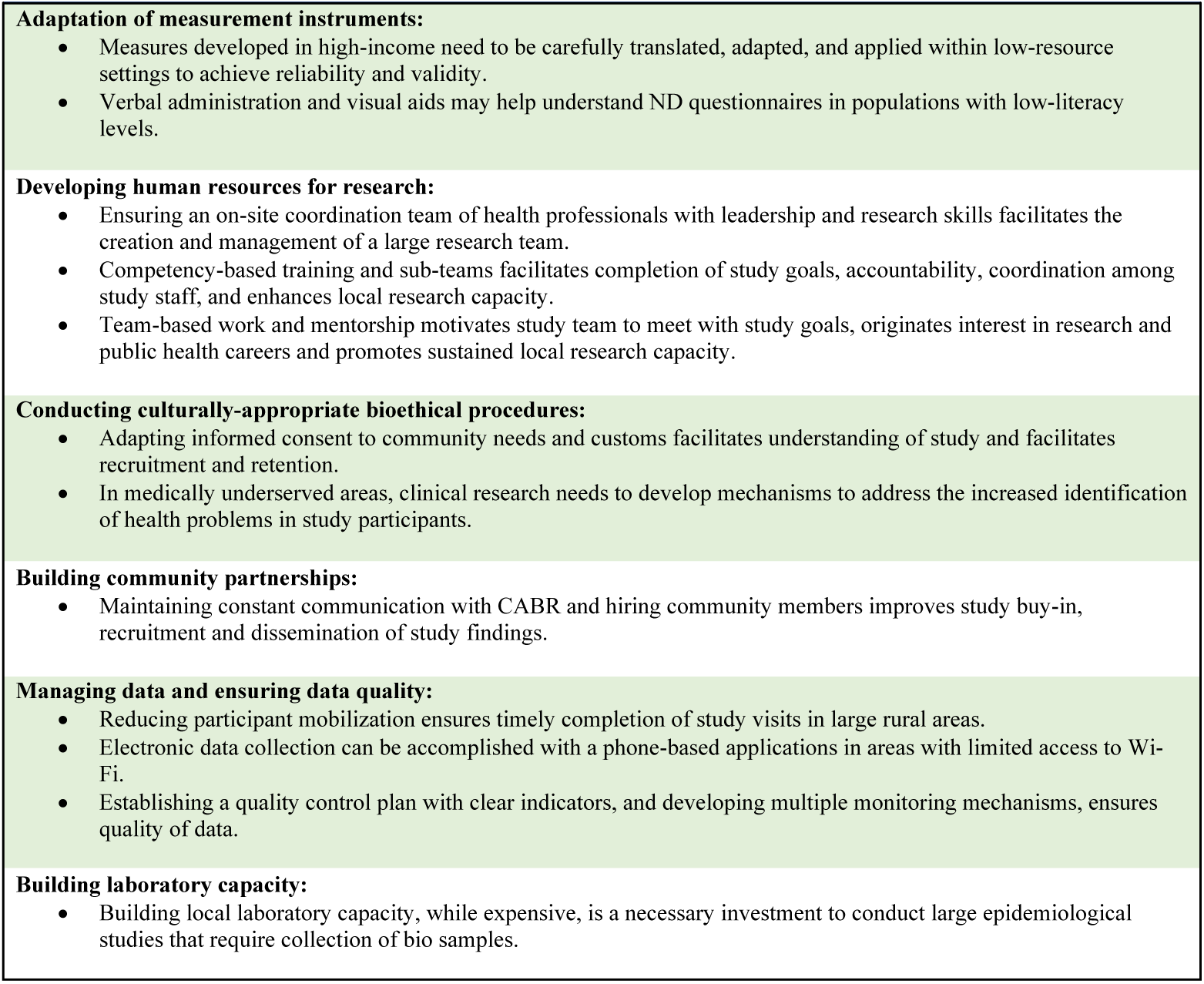
Lessons learned / Recommendations.

#### Adapting Instruments for Neurodevelopment Assessments

While many options for neurodevelopmental assessment exist in higher-income countries, few tests have been standardized and validated for use in low resource areas, and none for the Guatemalan population and our study population age range. ^15,16^ As this study aimed to evaluate children while the ZIKV epidemic was ongoing, the development of a new neurodevelopmental measure, which can take several years, was deemed impractical. ^17–19^ The process of the selection, translation and adaptation, and use of neurodevelopmental measures for this study has been previously described in detail. ^20^ In brief, a multi-disciplinary team composed of psychologists with content, cultural, and local expertise, along with neuropsychologists from the UC, selected the most appropriate test from existing tools, considering the relevance of these developmental domains in the local community and the developmental domains targeted by the study. We selected the Mullen Scales of Early Learning (MSEL) as our primary performance-based developmental assessment measure, and two parent report measures: the Ages and Stages Questionnaire-Third Edition (ASQ3), a developmental screening tool, and the Pediatric Quality of Life Questionnaire (PedsQL), a measure of health-related quality of life. Visual Likert scales were developed to aid parents’ understanding of questions and responses. Questionnaires were administered verbally to all parents to account for differences in levels of literacy and limited familiarity with neurodevelopmental assessments. Assessment of interrater reliability, internal consistency, and construct validity of the adapted and translated version of the MSEL was conducted as previously reported. ^21,22^ Psychologists were trained in how to maintain standardization conducting assessments in a variety of settings, including our clinic, a converted bus, satellite clinics, and in the home. Data analysis was done and demonstrated no significant differences in child performance between testing spaces.

#### Training and Managing Human Resources

Upon contract approval by NIH in January 2017, and due to the urgency to enroll participants in the study given the ongoing ZIKV epidemic, we proceeded to hire and train the study coordinator, three psychologists and one physician, focusing on personnel with expressed interest in research and public health. Two experienced research nurses, already hired by CHD became the field coordinators, and we rapidly expanded our team from 4 to 20 study nurses and three new data managers. All personnel were trained in NIH human subjects protection (HSP), Good Clinical Practices (GCP) and principles of clinical research following the NIH regulatory training, the Special Programme for Research and Training in Tropical Diseases (TDR) Global Competency Framework for Clinical Research, and the WHO Ethics in epidemics, emergencies and disasters: Research, surveillance and patient care manual.^23,24^

The study manual of procedures (MOP) and various standard operation procedures (SOPs) were developed, reviewed and refined as needed during the study operationalization. Study coordinators developed checklists summarizing visits and key procedures (e.g., enrollment visit, informed consent process, specimen collection and transport, data collection and electronic data entry) to train, supervise and provide feedback to the research staff on an ongoing basis. To ensure the implementation of multiple and complex procedures as required by the study protocol, we created competency-based sub-teams responsible for certain procedure clusters: recruitment and enrollment visits, symptomatic surveillance, FLI visits, and follow-up visits. Each sub-team included 2-6 study nurses and one field coordinator (**Figure 2)**. Weekly and daily specific and measurable goals were established and monitored by each field coordinator to ensure that enrollment, retention and study procedures goals were met.

Psychologists received on-site training from a CU neuropsychologist at the start of the study and then periodically throughout. Weekly videoconferencing, video reviews of assessments, and quality checks then occurred to ensure high quality neurodevelopmental assessments. As the number of study visits and procedures increased, we trained one group of nurses on physical examination and another group to conduct ASQ3 and PedsQL assessments with supervision by physicians and psychologists. This division of labor allowed us to complete procedures in a rapidly growing number of study subjects (**Figure 3)** within the required visit timeframes, while psychologist and physicians focused on more complex procedures and complicated cases.

**Figure 3.**
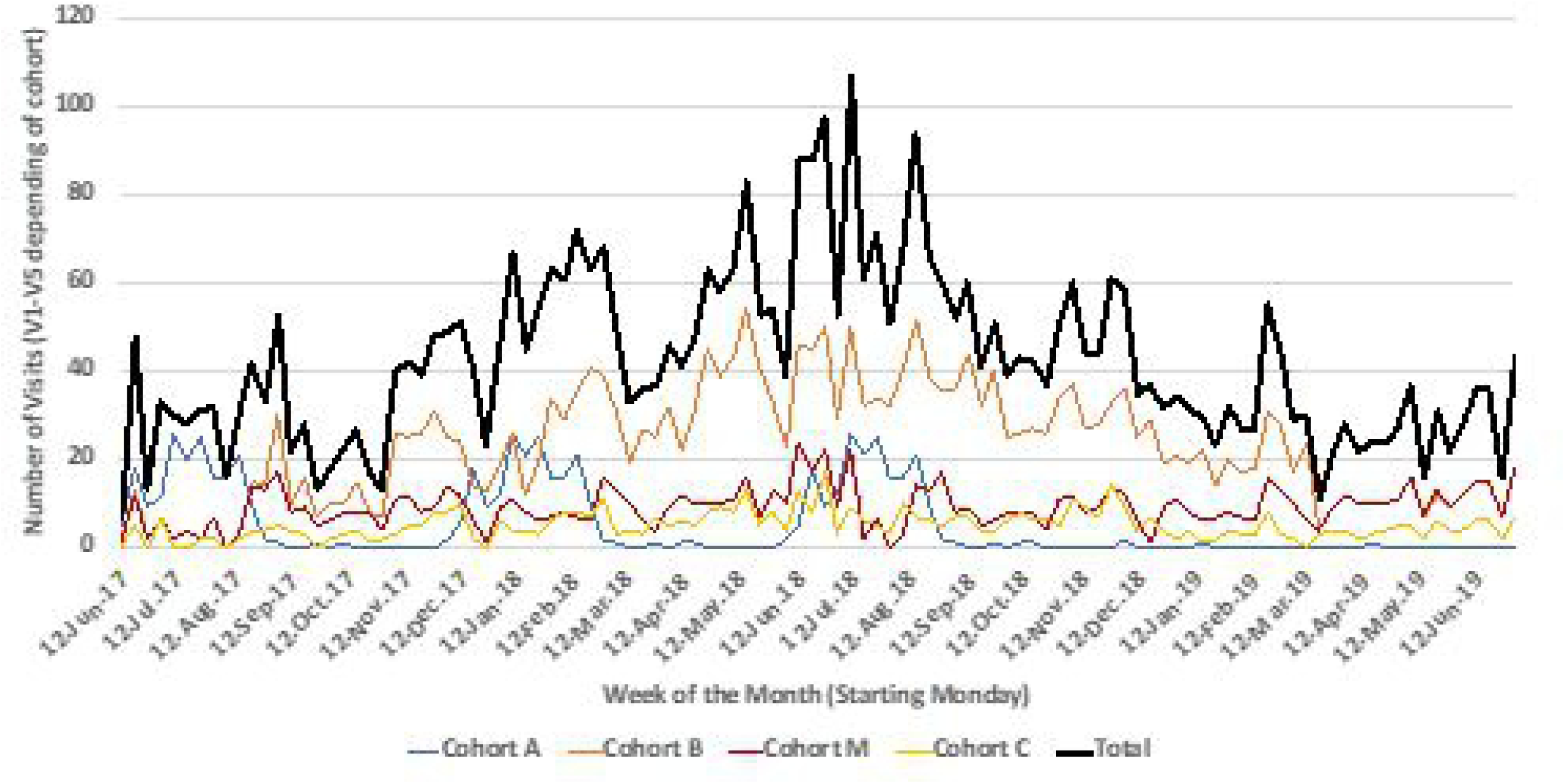
Enrollment and follow-up visits by study week.

A mentorship and training network with PIs, co-PIs and coordinators was established, to promote interest in research and public health among study staff. A strong culture of community service, continued learning and improvement, and team-based work was promoted throughout the study. This shared spirit ensured that study personnel stayed motivated to collaborate among sub-teams, and meet demanding study goals in a challenging environment.

#### Engaging Community Members

The community-based nature of this cohort study and the need to include villages in the study that were not familiar with CHD activities, required strong community engagement. During the 5-month preparatory phase, study coordinators and research nurses met with members of the local CABR and leaders of each of the 22 participating communities to explain the purpose and procedures of the study and to obtain feedback for its deployment. CHD established the CABR in 2015 to engage the local communities in all research and programmatic activities. The board is composed of community and religious leaders, teachers, and public health-post nurses who were trained by a consultant from the Colorado Multiple

Institutional Review Board (COMIRB) on how to review research studies and advocate for their communities. Research nurses from the participating communities provided their local expertise and helped enhance the study buy-in. Local leaders, midwives and nurses identified and referred potential study participants and maintained communication with families enrolled in the study. Community collaborators were trained on the essentials of GCP and Human Subjects Protection by an anthropologist from CU.

#### Adapting bioethical procedures to local context

Communities in SWT are characterized by involving community leaders and families in decision making. For this reason, it was important to socialize the study informed consent to communities and families, before individuals’ consenting. Prior to initiating research activities, lead study staff obtained community buy-in by presenting and discussing the study aims, procedures, and potential risks and benefits with community leaders. A simple and culturally-appropriate visual informed consent tool (Supplementary Appendix) was used to explain the study objectives and procedures in Spanish (the local language) to potential participants, highlighting the differences between research and healthcare services to families with low literacy levels and limited or no previous exposure to medical research. Informed participants older than 18 years old provided informed consent prior to study procedures. If participating mothers were 16 to 17 years old, informed consent was obtained from one of their parents and assent was provided by the mother. If the participant providing consent was illiterate, a third party witnessed the consent process and signed the informed consent form, while the participant provided thumbprints.

Dissemination of final results (aggregated and individual results to participants) was planned in collaboration with the Community Advisory Board for Research (CABR), community leaders and local public health facilities.

#### Collecting data and ensuring quality control

The study catchment area covered a large geographic zone, with deteriorated roads and limited transportation. For this reason, reducing participant mobilization and finding alternative sites, other than CHD clinic, to conduct study visits was needed. Visits were therefore conducted at multiple settings, including satellite clinics, a converted bus and participant’s homes. In order to collect large amounts of data in a location with limited access to internet, an electronic, smartphone-based CommCare app (Dimagi, Cambridge, MA, USA) was developed. Study staff collected data online or offline and uploaded it to an electronic server in real-time or as soon as internet was available, at least once daily. Research nurses completed most questionnaires (e.g., sociodemographic, medical history, illness visits, etc.), while trained psychologists and physicians documented neurodevelopmental and medical evaluations into paper-based source documents and later entered in the CommCare mobile application. In addition, information related to clinical visits and hospitalizations since the previous study visit was collected and entered into a secure local electronic REDCap database (Vanderbilt University, TN, USA) if participants consented.

To ensure adherence to GCP standards, a quality control plan was developed with a priority list of indicators for quality assurance. The quality control coordinator reviewed all the informed consent forms and 10-20% of the individual study records, conducting a comparison audit of the paper-based forms and the electronic databases. We aimed for 100% compliance with informed consent indicators. As data were entered in real-time, all completed data collection forms were reviewed daily by study coordinators. In addition, The Emmes Company, LLC, a data management group, updated online process quality reports daily or weekly allowing local data managers to address data issues (e.g., missing or duplicated forms, missing or extreme values, unexpected specimen samples) on an ongoing basis. Site specific report logs were maintained in a secure file sharing Website that provided real-time, site-specific feedback to identify errors and/or omissions in data entry and to facilitate corrections with minimal time delay.

Study coordinators conducted staff refresher trainings every two months as well as monthly field supervision of study staff procedures and data collection, while study investigators conducted random spot-checks of field work and home visits. Feedback through on-the-spot discussion of supervisors’ findings, weekly staff meetings, and monthly written reports to all staff and external collaborators was done with the aim of ensuring high quality performance of study procedures.

During weekly meetings, study progress, challenges and solutions were discussed among leaders from the field team, CU, Baylor University, Emmes, and NIH representatives. In addition, principal investigators conducted monthly on-site supervisions.

#### Building laboratory capacity for biological sample collection and processing

Trained research nurses collected blood, saliva, urine and breast milk samples at the participants’ households or at the satellite clinics per study protocol (**Table 1)**. Given the high ambient temperature (25-38°C), long transit times and harsh environment at SWT, a strict protocol for ensuring quality collection of samples and maintaining cold chain in the field was developed. Whole blood was collected in EDTA tubes for ZIKV and DENV serologic and molecular testing and processed to obtain plasma for ZIKV serologic and molecular testing, and other congenital infections. Saliva was collected using a CultureSwab Collection and Transport System by brushing the participants’ cheek and soaking it with saliva for at least 20 seconds, and the sample was then mixed with 200 uL of Invitrogen (Thermo Fisher Scientific, Carlsbad, CA) nucleic acid stabilizer buffer. Urine and breast milk samples for ZIKV molecular testing were collected in sterile containers. All samples were transported in temperature-controlled portable coolers and delivered to the study laboratory for storage at 2-8°C until processing, and final storage in a −80°C freezer until shipment. Blood samples retained included whole blood and plasma aliquots. Samples were shipped periodically to the NIH sample repository (Fisher Bioservices, Maryland, USA) preserving the cold chain and following international guidelines for shipping biologic materials. Specimen analyses included IgM, IgM, neutralizing antibodies and PCR.

In 2015, the CHD began enhancing its research capacity by building a 500 square foot (sqf) research laboratory and 1500 sqf research operations area for personnel, data management, secure document storage and meetings, with the support of the CU Strategic Initiative for Research. While the CHD has a connection to the power grid and a high-capacity generator, the stability of energy supply was reinforced by two additional generators to safeguard samples stored at the laboratory. During the conduct of this study, a solar-based energy system was installed. The laboratory had the capacity to process and store (−20da° C and −80° C, as appropriate) approximately 10,000

## Discussion

We described the rapid, intensive, and successful operationalization of a unique population-based prospective cohort study in a low-resource setting in rural Guatemala during the emergence of ZIKV in the Americas.

Our team faced multiple challenges due to the fast-paced nature of our work, large amounts of complex study procedures, and a study setting in a large geographic region, with high poverty rates, low literacy levels, and limited access to healthcare. Most importantly, we dealt with the very limited research infrastructure and resources that characterizes countries like Guatemala, and especially rural regions like SWT, where this study was conducted^25^. Despite these challenges, we operationalized the study protocol by translating the study objectives into an implementation plan that outlined the key milestones, activities and inputs, considered the local needs and assets, and included risk management and troubleshooting mechanisms. ^26^ Our plan focused on the areas of adaptation of neurodevelopmental assessments, human resources, bioethical procedures, community engagement, data collection and quality control, and collection of biological samples. Just as important as the main goals of a study with vulnerable participants is how one goes about achieving such goals. From planning to implementation, our overarching priorities were to develop culturally-appropriate study procedures, to respect and collaborate with community members, and to build sustainable local research capacity, while ensuring that GCP and NIH standards of research were met.

Population-based studies with well-defined denominators, like our study, have the ability to provide incidence rates of emerging infectious diseases, their consequences and other relevant outcomes in low-resource settings. For example, this study can provide information on the incidence of microcephaly and neurodevelopmental outcomes, which are increasingly being documented in low-resource settings.^10,27^ In addition, our results will help untangle the possible associations and interactions between microcephaly and neurodevelopmental outcomes and health determinants like poverty, malnutrition, and recurrent illnesses and infections. In parallel to our study, two other studies were launched in the region: the Centers for Disease Control and Prevention conducted a facility-based observational study on the clinical and virological manifestations of postnatal ZIVK (PMID: 29813148)^28^, and the prospective pregnancy cohort study of Zika in Infants and Pregnancy (ZIP) in six endemic countries (Brazil, Colombia, Guatemala, Nicaragua, Puerto Rico and Peru), which included follow up of thee infants up to one year of age (PMID: 31391005).^29^ In contrast to our study, these projects focused on congenital Zika and recruited participants from healthcare facilities, rather than communities. A recently published systematic review of postnatal symptoms and neurological complications of postnatal

ZIVK highlighted the scarcity and quality of published data, calling for prospective studies to improve our understanding of the frequency of signs, symptoms and complications of ZIKV exposure, and to investigate potential effects of ZIKV on neurodevelopment ^30^.

As an example of developing culturally-appropriate study procedures, the neurodevelopmental assessments were selected from standardized measures in high-income countries, translated to Spanish and adapted to the local culture. Verbal administration of neurodevelopmental questionnaires and visual aids for Likert scales facilitated parental understanding of questions and addressed the local limited literacy levels. The inclusivity and adaptation of the informed consent process to ensure community engagement and participation allowed us to meet GCP and bioethics standards. ^20^ Developing strong community engagement, including participation of community leaders and other stakeholders, enabled us to accomplish an 85% retention of study participants despite the heavy participant burden of weekly home visits and twice weekly sample collections. Engagement of the community does not end with the successful operationalization and conduct of the study; it needs to be followed by a process of feedback as well as frequent dissemination of study results to community leaders and participants throughout the study. This ensures the trust of the community in the process of research, provides an avenue for disseminating results, and promotes community participation in the inquiry of problems that affect them.

Assembling a highly trained and committed group of local researchers under the mentorship of experienced investigators is key to rapidly operationalize and sustain a study of this magnitude in the context of multiple local challenging conditions. Developing clear SOPs, forming sub-teams with specific roles, conducting well-planned trainings with frequent refreshers, performing on-site supervision and real-time feedback, and establishing strong team motivation for continuing education and career development are paramount to maintaining data quality and meeting study recruitment goals.

Population-health studies provide a unique opportunity to build local capacity and empower local investigators. Our study has provided many young nurses, who gained experience from well-structured ethical and research procedures trainings, with professional growth and special expertise that can help them contribute to enhancing local and regional clinical and public health practice. Many of our study supervisors have now advanced their careers by pursuing additional studies and degrees in epidemiology and public health, and they have grown to serve as study coordinators, laboratory managers, and data management coordinators, providing a positive circle of growth in which they serve as role models for young people and potentially become future leaders in their communities.

## Data Availability

All relevant data are within the manuscript and its Supporting Information files.

## Acknowledgements

We thank all co-investigators, study team, advisors, sponsors, and partners, especially SWT community members and parents whose support made the study possible. We want to thank especially Dr. Walla Dempsey, Mary Smith, Gail Tauscher, Dr. Lisreina Toro, John Brett, Dr. Kathryn Colborn, Cristina del Hoyo, Agroamerica, and all the study and clinic staff at FUNSALUD.

## Related manuscripts

No related manuscript is under consideration or accepted for publication elsewhere.

## Availability of Data and Materials

The protocol (version 1.0, January 23, 2017) for this study and the de-identified data from this study will be made publicly available on the NIAID ITN TrialShare platform in mid 2020.

